# Framework for prioritizing variants of unknown significance from clinical genetic testing in kidney disease – utility of multidisciplinary approach to gather evidence of pathogenicity for Hepatocyte Nuclear Factor-1β (*HNF1B*) p.Arg303His

**DOI:** 10.1101/2022.04.01.22273321

**Authors:** Uyenlinh L. Mirshahi, Ahana Bhan, Lotte E. Tholen, Brian Fang, Guoli Chen, Bryn Moore, Adam Cook, Prince Mohan, Kashyap Patel, Peter Igarashi, Jeroen H.F. de Baaij, Silvia Ferrè, Joost G.J. Hoenderop, David J. Carey, Alexander R. Chang

## Abstract

Monogenic causes in over 300 kidney-associated genes account for roughly 12% of end stage kidney disease (ESKD) cases. Advances in next generation sequencing, and large customized panels enable the diagnosis of monogenic kidney disease noninvasively at relatively low cost, allowing for more precise management for patients and their families. A major challenge is interpreting rare variants, many of which are classified as variants of unknown significance (VUS). We present a framework in which we thoroughly evaluated and provided evidence of pathogenicity for *HNF1B*-p.Arg303His, a VUS returned from clinical genetic testing for a kidney transplant candidate. This blueprint, designed by a multi-disciplinary team of clinicians, molecular biologists, and diagnostic geneticists, includes using a health system-based cohort with genetic and clinical information to perform deep phenotyping of VUS carriers, examination of existing genetic databases, as well as functional testing. With our approach, we demonstrate evidence for pathogenicity for *HNF1B*-p.Arg303His by showing similar burden of kidney manifestations in this variant to known *HNF1B* pathogenic variants, and greater burden compared to non-carriers. Determination of a molecular diagnosis for the example family allows for proper surveillance and management of *HNF1B*-related manifestations such as kidney disease, diabetes, and hypomagnesemia with important implications for safe living-related kidney donation. The candidate gene-variant pair also allows for clinical biomarker testing for aberrations of linked pathways. This working model may be applicable other diseases of genetic etiology.

## Introduction

Monogenic causes account for up to 40% of patients in cohorts enriched with end stage kidney disease (ESKD) or familial kidney disease (1-4). Genetic testing has become an invaluable tool in the diagnosis of various types of chronic kidney disease (CKD), including but not limited to autosomal dominant tubulointerstitial kidney disease (ADTKD), congenital anomalies of the kidney and urinary tract (CAKUT), focal segmental glomerulosclerosis (FSGS), and CKD of unknown cause (2, 5). Genetic testing allows for more precise molecular diagnosis—in some cases diagnostic reclassification. Benefits of establishing a molecular diagnosis include earlier treatment with disease-modifying therapies, screening for extrarenal manifestations, avoidance of invasive procedures such as kidney biopsy, and important implications for family planning and living kidney donation (3, 5, 6).

*HNF1B* encodes the hepatocyte nuclear factor 1β (HNF1B) and is a member of the homeodomain-containing superfamily of transcription factors involved in the development of kidney, urogenital tract, pancreas, liver, brain, and parathyroid gland (2). Pathogenic *HNF1B* variants lead to a wide spectrum of phenotypic expression ranging from non-insulin dependent, maturity onset diabetes of the young (MODY), pancreatic hypoplasia, liver cholestasis, and renal cysts, renal agenesis, and collecting system abnormalities, hypomagnesemia, hypermagnesuria, hyperuricemia and hyperparathyroidism (7-10). The phenotypic expression of *HNF1B* carriers varies even between individuals with the same mutation within families, possibly as a result of temporal stochastic variations in *HNF1B* expression during nephrogenesis(9). As there are a large number of genes associated with *HNF1B* renal phenotypes (ADTKD, CAKUT, cystic kidney disease), the use of exome sequencing or massively parallel sequencing (MPS) can be very helpful (1, 2, 4, 5, 11-13).

MPS has become increasingly available and more affordable in clinical practice(14, 15). MPS allows the evaluation of multiple genes and thus can be particularly useful for monogenic kidney disorders with broad differential genetic causes. A consequence of testing dozens if not hundreds of genes linked to kidney disease is that many variants of unknown significance (VUS) are often detected. In various cohorts, up to 10-30% of genetic results may be VUS, classified according to the American College of Medical Genetics-Association for Molecular Pathology (ACMG-AMP) criteria (3, 16-18). VUS present a diagnostic and ethical challenge in genetic testing and lack of resolution may result in delays in treatment and management (19). Variant reclassification from VUS to (likely) benign, or (likely) pathogenic improves as data sharing and variant curation efforts from expert panels (e.g. ClinGen and Genomics England) expand (5). Apart from case studies, implementation and strategies to efficiently transition kidney VUS to a more definitive classification are lacking.

Clinically unselected research population databases present a unique resource that can be used to triage VUS. The MyCode DiscovEHR database, currently comprised of over 173,000 individuals who have exome sequencing and linked-electronic health records (EHR) (20), includes rich, longitudinal in-patient and out-patient data on multiple generations spanning over 20 years that could be leveraged for studying clinical features of carriers of VUS that are returned from clinical genetic testing.

In this exemplar study, we present a framework of efforts from a multidisciplinary team with expertise in nephrology, endocrinology, molecular biology, and diagnostic genetics to gather evidence of pathogenicity for a VUS, *HNF1B* c.907C>T p.Arg303His, identified through a clinical MPS panel for a patient with CKD of unknown cause.

## Methods

### Study populations, genetic testing, clinical data abstraction

#### Clinical family

The index patient (proband) was referred to Geisinger Medical Center for kidney transplant evaluation. The patient and additional family members who were potential kidney donors were referred for clinical genetic testing. A genetic testing panel was performed using Natera Renasight for exonic and copy number variations (CNV) in 385 genes associated with kidney disease (Natera, TX). Pertinent lab testing (serum magnesium, fractional excretion of magnesium (FEMg), uric acid, renal and liver function tests) were offered as part of routine care. Renal and extra-renal involvement was recorded by patient interview and review of electronic health records (EHR). Permission to publish the case series was obtained from the proband and her family members.

#### Geisinger DiscovEHR cohort

The Geisinger DiscovEHR cohort consisted of 172,589 individuals who received healthcare at Geisinger, a health system in central and north-eastern Pennsylvania, USA. Individuals consented to participate in the MyCode Community Initiative to create a biorepository of blood, serum and DNA samples for broad research use, including genomic analysis(20), with linkage to the EHR. Renal and extra-renal involvement in the cases including clinical diagnosis, procedures, imaging results, and laboratory results were reviewed. Genetic analysis was carried out as part of the DiscovEHR collaboration between Geisinger and the Regeneron Genetics Center by exome sequencing (21). This study was reviewed by the Geisinger Institutional Review Board and determined as not including human subject research as defined in 45CFR46.102(f) in written consent (Study # 2021-0177).

#### Exeter cohort

The Exeter cohort comprised of >5000 individuals with clinical suspicion of monogenic diabetes referred from routine clinical practice across the UK for genetic testing at the Molecular Genomics Laboratory at the Royal Devon and Exeter Hospital, Exeter, UK. Informed consent was obtained from these patients or their parents/guardians. The study was approved by the North Wales ethics committee.

### Variant annotation and classification

The variants from clinical genetic testing and from Geisinger DiscovEHR research studies were annotated using clinically relevant RefSeq transcripts and classified by the clinical genetic laboratories or at Geisinger (DiscovEHR study) using ACMG-AMP guidelines (22, 23). *HNF1B* copy number loss were called from a referenced-based CLAMMS algorithm using exome sequencing (24), and confirmed by Illumina chip array. Samples underwent quality assurance including removal of samples of sex mismatch, other large chromosomal abnormalities, and outliers of derivative log ratio spread (DLRS) and genomic wave factors (25). Outliers were defined as 1.5 times the interquartile range (IQR) from the third quartile of the distribution of DLRS. Patients referred as having a *HNF1B* whole gene deletion in this study included 25 clean samples with 17q12 microdeletion, a 1.26Mb deletion ranging from *AATF* to *HNF1B* which results in complete loss of the entire *HNF-1B*. Noncarriers in this study refers to samples in the MyCode cohort without 17q12 microdeletion, known pathogenic *HNF1B* variants as defined by ClinVar(26), or protein truncating variants.

### Luciferase reporter assay

Human embryonic kidney 293 (HEK293) cells were seeded in 24 well plates and transfected with 350 ng of the promoter firefly luciferase constructs (pGL3) previously generated containing the kidney-specific promoters of Na^+^/K^+^-ATPase subunit gamma (*FXYD2*), or Polycystic Kidney and Hepatic Disease 1 *(PKHD1)*, or empty vector (27). All three constructs were co-transfected with 25 ng of pCINEO-empty, pCINEO-*hHNF-1B*, pCINEO-*hHNF-1B*-Arg303His or pCINEO-*hHNF-1B*-Lys156Glu cDNAs. Additionally, 10 ng of Renilla luciferase construct (pRL) under a CMV promoter was co-transfected and used as a control the transfection efficiency. Co-transfections were performed using polyethylenimine cationic polymer (PEI) (Thermo FisherScientific) in 1:6 DNA to PEI ratio. For dose-response experiments, HEK293 cells were co-transfected with human *FXYD2 or PKHD1* promoter construct and 50 (PKHD1 only), 25, 12.5, 6.25, or 3.125ng pCINEO-*hHNF1B* or pCINEO-*hHNF-1B*-Arg303His cDNAs. Firefly and Renilla luciferase luminescence were measured 48 hours after transfection with the dual-luciferase reporter assay (Promega) using a plate reader (VICTOR, PerkinElmer).

### Biomarkers

We compared presence of clinical labs (eGFR, magnesium, amylase, lipase, uric acid) related to *HNF-1B*-related disease among *HNF1B*-Arg303His heterozygotes to individuals carrying the 17q12 microdeletion encompassing *HNF1B*, and non-carriers. Estimated GFR was calculated for individuals age <18 years using the Schwartz equation (28), and age > 18 years using the CKD-EPI equation (29). Longitudinal outpatient serum biomarkers were restricted to the age range of the *HNF1B*-p,Arg303His cases (15-60 years); measures following kidney transplantation and dialysis were excluded. Measures from each patient were pooled per group, regressed using a linear model against age at measurement, and plotted with 95% confidence intervals (95% CI) using Matplotlib seaborn library. Regression statistics were generated using R lm package.

## RESULTS

### Kidney disease is observed in 3 generations of Family A in carriers of HNF-1B-p.Arg303His

Family A Proband, *Case AIII-1* is a 25-30-year -old female with a history of chronic pancreatitis, chronic kidney disease (CKD) stage 4, and asthma presented for kidney transplant evaluation. Between the ages of 10-15, she had been evaluated for elevated amylase and lipase, abdominal pain, CKD with no blood/urine on urinalysis. A kidney biopsy showed chronic tubulointerstitial nephritis and oligomeganephronia, and imaging showed diffusely hyperechogenic, small kidneys (8.8cm-left, 9.2cm-right) (Figure 1-A-C). Clinical genetic testing for hereditary pancreatitis (Ambry Genetics panel for *CFTR, SPINK1, PRSS1*) showed that she is heterozygous for *CFTR*-p.Leu1065Pro, a pathogenic variant. Evaluation of sweat test, and pulmonary and genetics consultation eliminated the variant as causal for her pancreatic issues. Although her BMI was low-normal (21.5 kg/m^2^), no testing for pancreatic insufficiency had been performed at that time. Between ages of 10-15 years, allopurinol therapy was initiated as she was found to have an elevated uric acid level (7.7 mg/dL). Between the ages of 20-25, her CKD had progressed along with proteinuria (511 mg/g protein/creatinine ratio). A repeat kidney biopsy done between the ages of 20-25 years showed progressive chronic tubulointerstitial injury, moderate/patchy interstitial fibrosis and tubular atrophy, mild fibro-intimal thickening of arteries, and oligomeganephronia (Figure 1). Further testing revealed chronically elevated amylase/lipase, with normal liver function tests (Table 1). Over several years, serum magnesium levels ranged from 1.4-1.8 mg/dL, and between 20-25 years old her serum magnesium was 1.9 mg/dL with FEMg indicating magnesium wasting (15%). Repeat abdomen/pelvis non-contrast CT between the ages of 20-25 years showed small kidneys and an unremarkable liver and pancreas.

**Figure 1.**
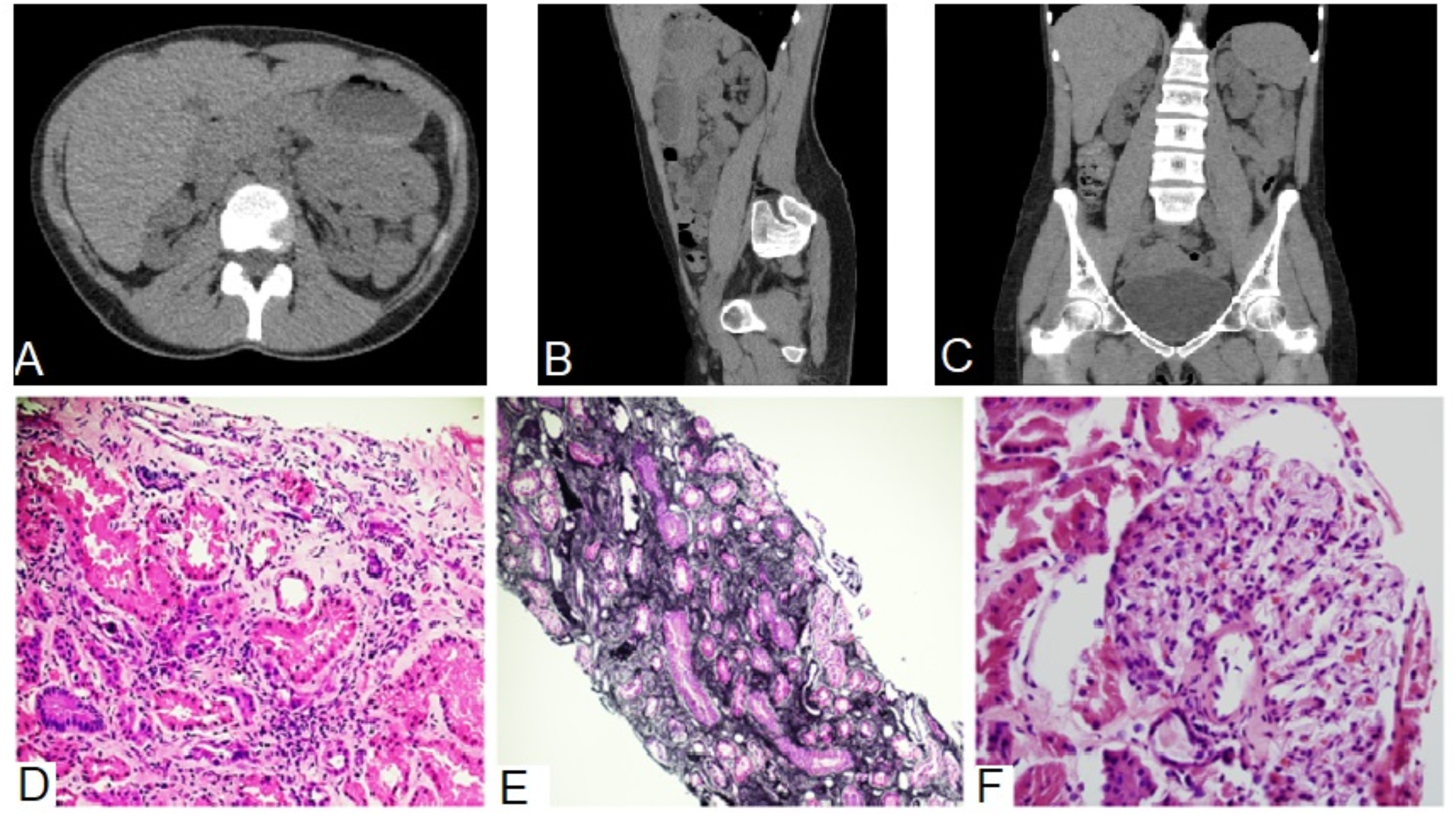
Imaging and histopathology of Family A Proband (Case AIII-1) *HNF1B-*p.Arg303His carrier showed bilateral small kidneys and oligomeganephronia. Axial (A), sagittal (B), and coronal (C) images from non-contrast CT scan taken at age between 25-30 years show small kidneys without cysts, and no overt abnormality of the pancreas and the liver. D) Hematoxylin and eosin (H&E) staining of renal biopsy specimen showed focal interstitial inflammation and mild tubular injury (10×10 magnification). Focal interstitial fibrosis is also present, which is highlighted by Jones silver staining in E) (4×10 magnification). F) In the cortical areas, the number of glomeruli is reduced. Mild tubular hypertrophy associated with mildly enlarged glomeruli is observed as shown in a representative image (20×10 magnification).

**Table 1.** Clinical characteristics of heterozygous carriers of *HNF1B* c.908G>A p.Arg303His at first presentation.

| Characteristics | Family A |  |  | Family B | Family C |  |
| --- | --- | --- | --- | --- | --- | --- |
|  | Case AIII-1<br>(Index Case) | Case AII-1 | Case AIII-2 | Case BI-1 | Case CII-1<br>proband | Case CI-1<br>Proband's parent |
| Study | This study | This study | This study | This study | Snoek *et al.* | Snoek *et al.* |
| Age, years | 25-30 | 45-50 | 20-25 | 55-60 | 30-35 | 45-50 |
| Sex | Female | Female | Female | Male | Male | Female |
| BMI, kg/m<sup>2</sup> | 21.5 | 19.8 | NA | 25.9 | NA | NA |
| Hypertension | No | No | No | No | NA | NA |
| Diabetes (age) | No | No | No | Yes (51) | NA | NA |
| ESRD (age) | Yes (25) | No | No | No | NA | NA |
| Genitourinary abnormalities | No | Ureteral stenosis requiring dilatation in her 20s | No | No | NA | NA |
| Pancreas Abnormalities | Pancreatitis | Pancreatitis | No | Mild atrophy Pancreatitis | NA | NA |
| Creatinine, mg/dL | 3.2 | 1.6 | 1.2 | 1.7 (50s) | NA | NA |
| eGFR ml/min/1.73m<sup>2</sup> | 20.7 | 39.0 | 62.7 | 45.6 (50s) | 39 (30s) | NA |
| Albuminuria or proteinuria | ACR: 331 mg/g | ACR: 37.3 mg/g | Dipstick: Trace | Dipstick: Negative | 24-hour urine protein: 0.6 g/day | NA |
| Microscopic hematuria | Negative | Negative | Negative | Negative |  | NA |
| Albumin g/dL | 4.5 | 4.4 | NA | NA | Normal | NA |
| AST U/L | 15 | 32 | 30 | 23 | Normal | NA |
| ALT U/L | 10 | 44 | 9 | 33 | Normal | NA |
| Alkaline phosphatase U/L | 51 | 59 | 220 | 73 | Normal | NA |
| Serum magnesium mg/dL | 1.9 | 1.7 | 1.8 | 1.6 (40s) | NA | NA |
| FEMg % | 15 | 9 | 3 | NA | NA | NA |
| Uric Acid, mg/dl | 7.7 (<18) | 5.8 (40s) | 4.9 (20s) | NA | NA | NA |
| Kidney biopsy findings | Chronic tubulointerstitial nephritis, oligomeganephronia at adolescent years | Zonal interstitial fibrosis thought to be due to ureteral stenosis in her 40s | NA | Chronic kidney disease, unknown etiology | FSGS unknown etiology with 45% glomerulosclerosis in her 30s | Hypodysplasia |
| Abdominal imaging | Small diffusely hyperechoic kidneys, normal pancreas on MRI at adolescent years<br>-Small kidneys, unremarkable pancreas by CT in her 20s | NA | NA | Kidney unremarkable, mild atrophy pancreas | Kidney unremarkable in appearance and size | NA |

Abbreviations: ESRD end-stage renal disease, eGFR estimated glomerular filtration rate, ACR (albumin/creatinine ratio), AST aspartate aminotransferase, ALT alanine aminotransferase, FEMg fractional excretion of magnesium, FSGS focal segmental glomerulosclerosis, CT computed tomography, MRI magnetic resonance imaging. If lab values were obtained at an earlier age, the age appears in parentheses.

### Additional family testing reveals co-segregation of the HNF-1B-p.Arg303His variant with disease in Family A

Clinical genetic testing performed using the 385-gene Natera Renasight panel revealed that the proband’s parent and sibling both harbored *HNF1B* Arg303His (Supplemental Table 1, Figure 2). Family A proband’s parent, *Case AII-1*, had a history of pancreatitis, chronically elevated amylase and lipase, and ureteral stenosis for which she underwent ureteral dilation in her late 20s (Table 1, Figure 2). Her BMI was low-normal at 19.8 kg/m^2^, and she had never undergone testing for pancreatic insufficiency. No history of diabetes or liver problems were noted. She developed CKD in her late 30s, (eGFR 43 between 45-50 years) with no proteinuria or hematuria. Magnesium between 35-40 years old was low at 1.4 mg/dL; FEMg at this time was 9%. Kidney biopsy revealed somewhat dilated tubules, mild arterial fibro-intimal thickening without active interstitial inflammation. No renal or pancreatic imaging was available for this patient. Family A proband’s younger sibling (between20-25 years old), *Case AIII-2*, had eGFR levels <1^st^ percentile for her age/sex ranging 63 and 67 mL/min/1.73m^2^ in the past year (Table 1, Figure 2). She has no diabetes or known abnormalities of the liver or pancreas although she had not received testing for pancreatic insufficiency or abdominal imaging. FEMg measured between the ages of age 20-15 was 3%. Family A proband’s maternal great relative (*Case AI-2*, Figure 2) had a history of hypertension and kidney disease at 20-25 years old and was born with one of her kidneys being smaller than the other. Per the proband’s paremt, Case AI-2 later had kidney failure and died in her 50s. No additional clinical information is available. Family A proband’s grandparent (*Case AI-4*, Figure 2) was reported to have a possible history of pancreatitis.

**Figure 2.**
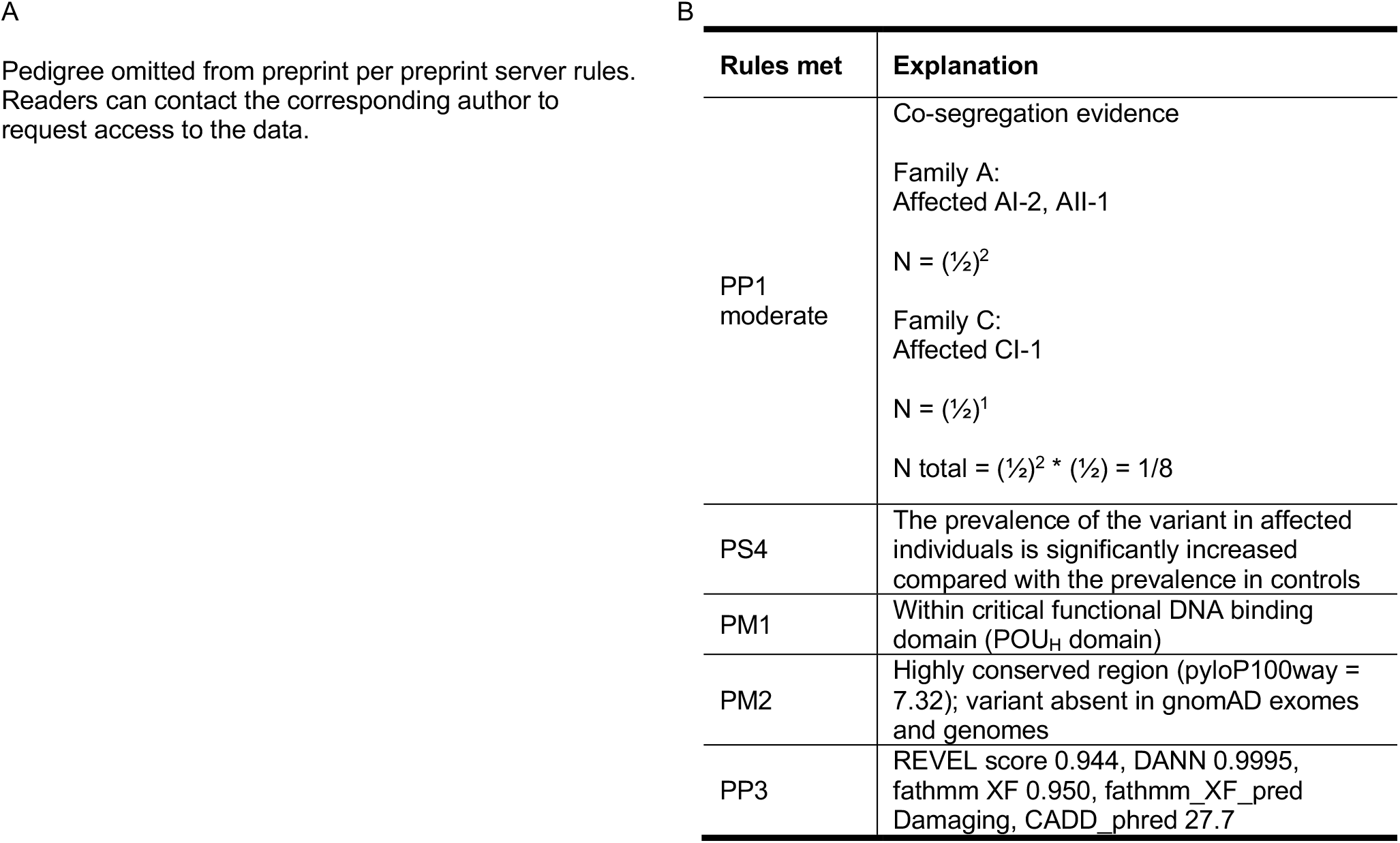
Family A pedigree and pathogenic classification of *HNF1B-*p.Arg303His. **A.** Male members are represented in squares, females in circles. Filled-in symbols indicate affected members with clinical features previously observed in *HNF1B* loss of function patients are shown, open symbols show unaffected members, and hashed symbols are members highly suspicious for the kidney disease. A positive sign indicates that the individual is heterozygous for *HNF1B-*p.Arg303His, and a negative sign indicates that the individual was genotyped and does not harbor Arg303His. The proband is identified with an arrow. Each generation with informative health records are labeled with Roman numerals on the left. **B**. Pathogenic classification for *HNF1B-*p.Arg303His per ACMG-AMP guidelines. The rules and the explanation for meeting the rules are indicated. For co-segregation evidence, see text. Abbreviations: HTN hypertension, CKD chronic kidney disease, eGFR estimated glomerular filtration rate

Genetic testing of unaffected family members (proband’s maternal relative (*Case AII-2*, Figure 2) and maternal cousin (*Case AIII-3*) as potential kidney donors using the same MPS panel test found that neither unaffected individual carried the *HNF1B* Arg303His. Incidentally, Case AIII-3 was found to have a likely pathogenic frameshift in *CFI* (c.1311dup p.Asp438Argfs*8).

### The MyCode population contains one participant carrier of HNF-1B-p.Arg303His who also has had CKD and pancreatitis

We interrogated the MyCode cohort of 172,589 participants and identified only one individual with *HNF1B* p.Arg303 and no other kidney associated pathogenic genetic variants. This man in his late 50s (Family B, *Case BI-1*), who harbored a p.Arg303His (minor allele frequency 2.9e-6), has clinical features of *HNF1B* mutations including a history of gout, elevated amylase, lipase and pancreatitis, and diabetes with insulin treatment for at least 10 years. Between the ages of 50-55, he has CKD Stage 3 (eGFR 45.6 mL/min/1.73m^2^, Table 1). Last CT taken between 50-55 years indicated mild atrophy of the pancreas. No cysts were observed in the liver or kidneys, and no genitourinary defects were noted.

### Review of genetic databases and literature

The *HNF-1B*-p.Arg303His variant is absent from the Broad gnomAD dataset. In reviewing the literature, a case was submitted to Clinvar (https://www.ncbi.nlm.nih.gov/clinvar/RCV001253232/) in which an individual was reported with RCAD; no other information was noted. A case study looking at genetic causes of FSGS reported a parent-child pair with ESKD at a young age who were heterozygous for *HNF1B*-p.Arg303His (30). The proband had ESKD between the ages of 30-33 (Family C, *Case CII-1*, Table 1) and FSGS on kidney biopsy. No renal structural abnormalities, electrolyte, glucose, or liver enzyme abnormalities were observed. The parent (*CI-1*, Table 1) was reported to be a carrier of the same variant and deceased at age 50 with ESKD, presumably from hypodysplastic kidneys.

### Serum biomarkers of *HNF1B*-p.Arg303His cases indicate decline in kidney and pancreatic function, similar to *HNF1B* whole gene deletion

As no differences in renal and extrarenal clinical characteristics between patients with *HNF1B* coding and splicing pathogenic variants and those with *HNF1B* whole gene deletion (17q12 microdeletion) have been reported (9), we compared eGFR, magnesium, and pancreatic enzyme levels of *HNF1B*-p.Arg303His cases to individuals with 17q12 microdeletion and noncarriers in the MyCode cohort. Figure 3 shows outpatient lab measures of cases in Family A and Family B compared to the lifetime profile of carriers with 17q12 microdeletion and noncarriers of pathogenic *HNF1B* variants. As expected, eGFR decline over time for 17q12 carriers were greater (−1.7 ml/min/1.73m^2^/year [95% CI -1.9, -1.4]) vs. noncarriers (−0.928 ml/min/1.73m^2^/year [-0.932, -0.924]; p value <0.0001). All *HNF-1B*-p.Arg303His carriers had eGFR levels lower than noncarriers and 17q12 carriers at the same age (Figure 3). Serum magnesium in all *HNF-1B*-p.Arg303His cases were similar to levels from 17q12 microdeletion carriers and lower than noncarriers (Figure 3). Three of the four *HNF-1B*-p.Arg303His cases had elevations in lipase whereas 17q12 microdeletion carriers had no differences in mean lipase or amylase levels compared to noncarriers.

**Figure 3.**
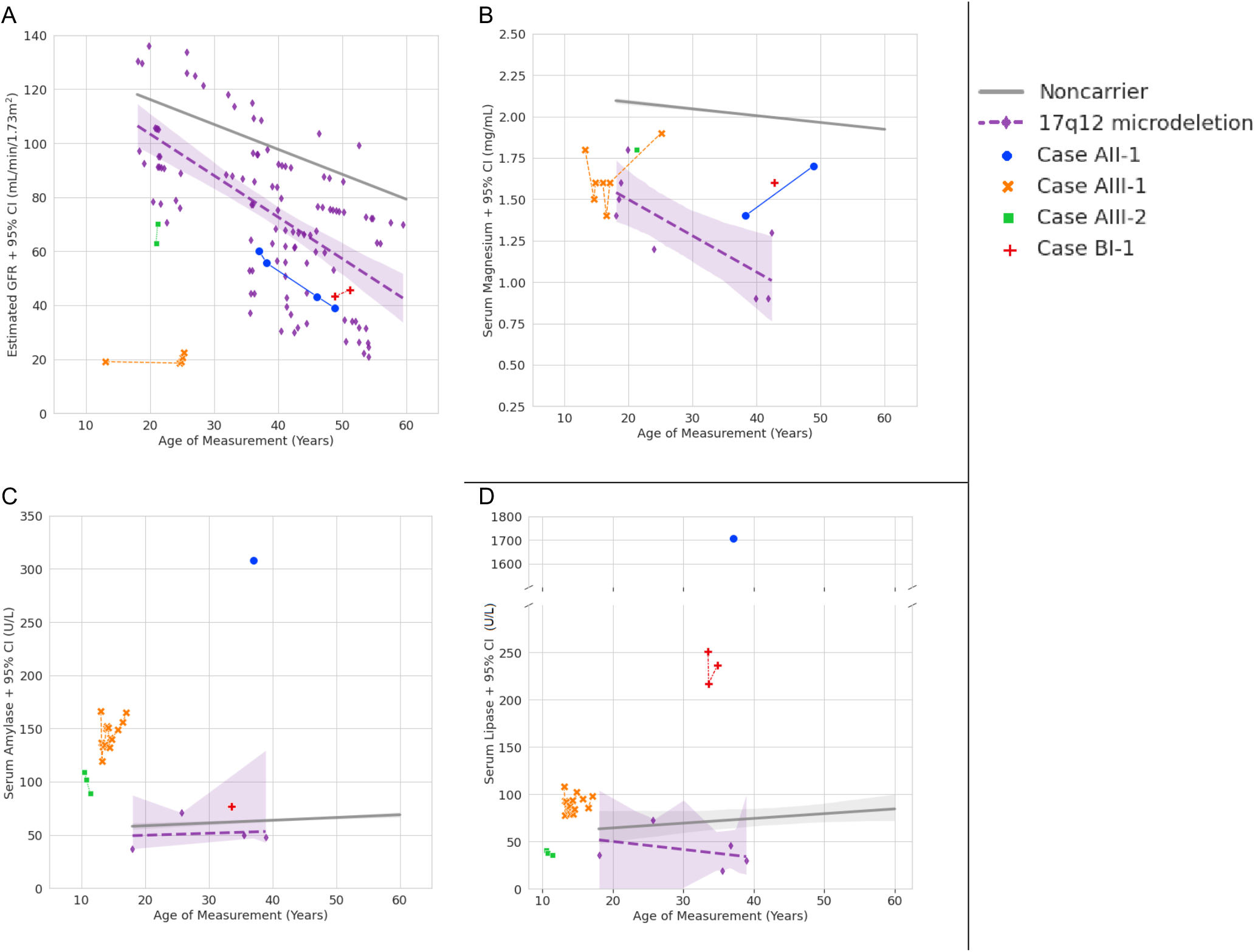
Serum biomarkers of *HNF1B-*p.Arg303His cases indicate decline in kidney and pancreatic function similar to *HNF1B* whole gene deletion. Lifetime lab measures from age 18-60 years for individuals with *HNF1B* whole gene deletion (17q12 microdeletion), *HNF1B*-p.Arg303His (Case AII-1, Case AIII-1, Case AIII-2, and Case BI-1), and noncarriers of *HNF1B* are plotted against the age of measurement. Linear regression was performed for 17q12 microdeletion and noncarrier groups. For clarity, only regression lines for noncarriers and 95% confidence intervals (95% CI) are shown. Data for *HNF1B*-p.Arg303His are shown as individual measurement per case due to a smaller number of tests. Biochemical measures and number of individuals with measures are as follow: A. Estimated GFR (n for noncarriers=115,298; n for 17q12 microdeletion=12), B. Serum magnesium (n noncarriers=21,411; n 17q12 microdeletion=4), C. Serum lipase (n noncarriers=24,323, n 17q12 microdeletion=4), D. Serum amylase (n noncarriers=18,116; n 17q12 microdeletion=4). See materials and methods for details.

### Carriers of other variations at the Arg303 locus also show renal abnormalities consistent with pathogenic *HNF1B* variants

To our knowledge, other *HNF1B*-p.Arg303 variations have been observed in only 3 other individuals to date. Laliève *et al*. reported a school-aged heterozygous carrier of *HNF1B* c.907C>T p.Arg303Cys with multi-cystic dysplasia (LOVD Patient ID 00231109, http://www.lovd.nl/HNF1B). In addition, from the Molecular Genomics Laboratory at the Royal Devon and Exeter Hospital, we report a parent-child pair with diabetes, both of whom harbor c.907C>A p.Arg303Ser. The child was referred for genetic testing for *HNF1B* due to early-onset diabetes (mid 20s) and solitary kidney. The parent had diabetes in her late 30s and also had the same variant. Diabetes was also present in five maternal uncles and aunts but genetic analyses on these family members are not available.

### *HNF1B* Arg303His mutation mildly affects *FXYD2* and *PKHD1* promoter activation *in vitro*

*HNF1B* acts as a transcriptional activator or repressor by binding to the promoter of target genes. We tested the binding properties of Arg303His on two kidney-specific promoters, *PKHD1* or *FXYD2*, in a dual-luciferase reporter assay system. *HNF1B*-p.Arg303His transcriptional activity on *FXYD2* promoter significantly increased by 13% compared to wildtype *HNF1B (*Supplemental Figure 1A); however, saturation curves with decreasing concentrations of promoter construct for *FXYD2* showed comparable transcriptional activity between the mutant Arg303His construct and wildtype *HNF1B* (Supplemental Figure 1B). In contrast, *HNF1B*-p.Arg303His showed a reduced transcriptional activity to wildtype *HNF1B* for *PKHD1* promoter (11%, p<0.05, Supplemental Figure 1A); however, the difference was not observed at saturating concentrations of 25 and 50 ng constructs (Supplemental Figure 1C). These studies suggest that compared to wildtype, the His at position 303 mildly alters transactivation potential of *HNF1B* in the tested *HNF1B* target promoters.

### *HNF-1B*-p.Arg303His classification per ACMG-AMP guideline

We combined all the evidence of *HNF1B*-p.Arg303His family members to classify *HNF1B*-p.Arg303His per ACMG-AMP criteria. Per Jarvick and Browning, we assumed that the variant has full penetrance and that it is inherited from one ancestor given its absence in gnomAD exomes and genomes and rare frequency in the MyCode population (MAF=2.9e-6) (23). By this assumption, for Family A, members AI-2 and AI-4 are untyped carriers (Figure 2A). Using a conservative approach, we considered AI-2 and AII-1 as affected carriers for Family A, and CI-1 as an affected carrier in Family C. We calculated the total N, the probability that the observed variant-kidney phenotype co-segregation is not by chance. The total N is the product of all Ns in each family. By definition, the probability of each proband in the three families is 1. We assigned N=½ for each affected carriers, resulting in N_total_ =1/8 (Figure 2B). A probability of 1/8 in two families provides a PP1 moderate level of pathogenicity support that the observed kidney phenotype did not occur by chance. Of note, the probability for co-segregation would have been higher had we included the contributions from the likely affected variant carriers (AI-4 and AIII-2) and the unaffected noncarrier members (AII-2 and AIII-3) in Family A. Per ACMG-AMP guidelines, additional supportive evidence confirming pathogenicity of this variant includes (Figure 2B): rarity of this variant in MyCode and its nonexistence in control populations like gnomAD; prevalence of the variant in affected individuals is significantly higher than in unaffected individuals; variant is in a hotspot of the gene that is highly conserved; *in silico* evidence suggests a deleterious effect on the gene product.

## DISCUSSION

In an effort to streamline the genetic diagnosis for a kidney transplant candidate, we present a framework to prioritize VUS results from clinical genetic testing for kidney disease developed through efforts from a multidisciplinary team (Figure 4). The workflow following receiving VUS results from clinical genetic testing consists of a literature review of all VUS for existing cases, variant analysis and deep phenotyping of carriers of all VUS in a large population cohort with to rule out unlikely VUS and provide evidence for a candidate VUS for *in vitro* functional testing. Additional clinical biomarker testing for clues of perturbation of biological pathways linked to the candidate gene-variant (e.g. FEMg) and co-segregation studies of kidney disease phenotype and variant carriers lead to adequate evidence to classify this variant as pathogenic per the ACMG-AMP guidelines and a genetic diagnosis of ADTKD-*HNF1B* in a patient with CKD of unknown origin.

**Figure 4.**
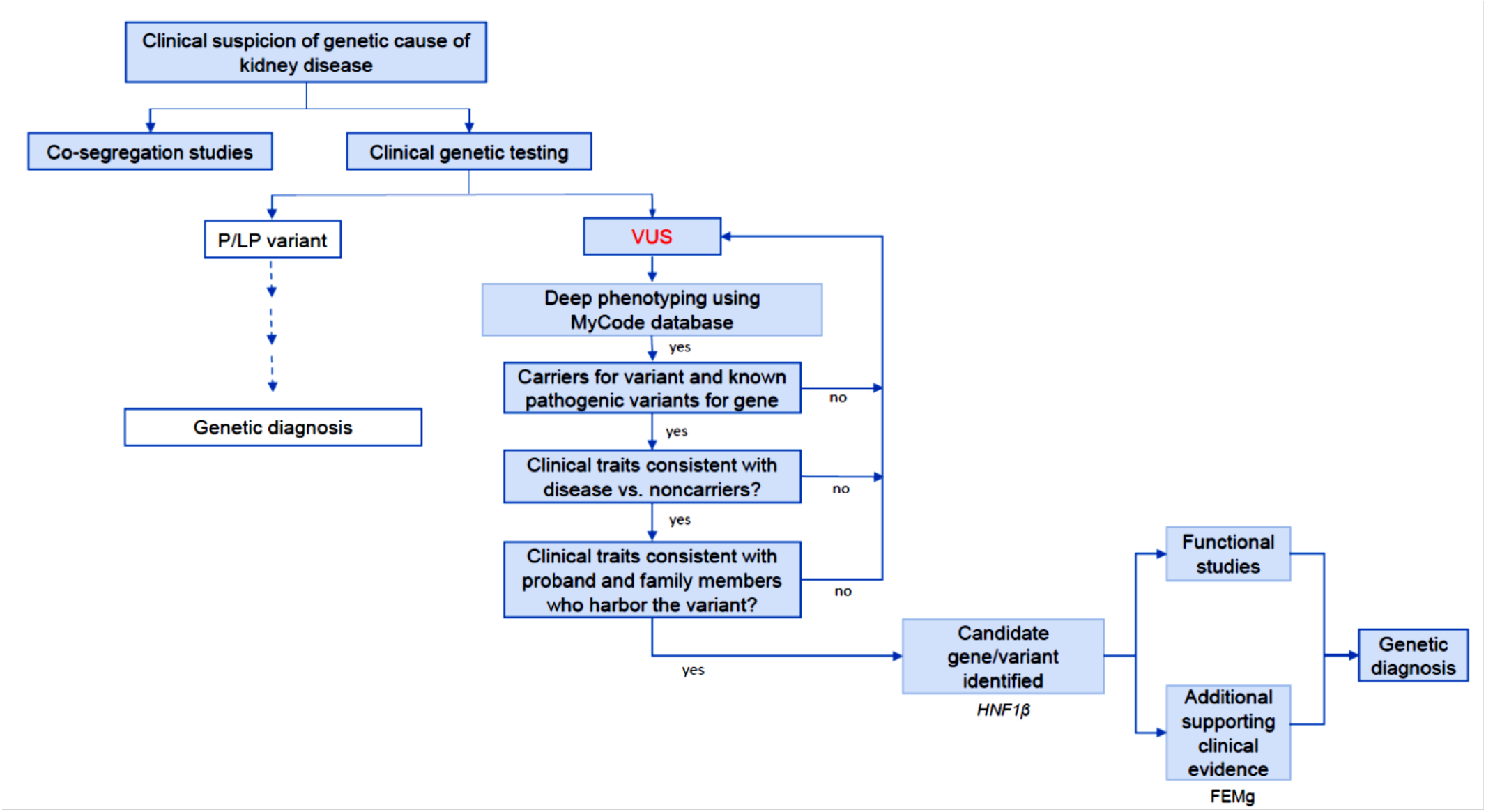
Framework for prioritizing VUS from clinical genetic testing. Clinical genetic testing returned multiple VUS for an kidney transplant candidate and her family members. Identifying the VUS and other known pathogenic variants of that gene in the MyCode database and deep phenotyping of carriers compared to noncarriers narrow the VUS pool to *HNF1B-*p.Arg303His. Studies of variant effect on protein function, additional clinical workup (Mg wasting), and studies of variant co-segregation with disease traits provide further supporting evidence of pathogenicity. Abbreviations: P/LP pathogenic/likely pathogenic, VUS variant of unknown significance, FEMg fractional excretion magnesium

Our index case and four other family members had multiple VUS returned from clinical genetic testing. There were only two VUS that were shared amongst the family members: *PTH1R*-p.Ala72Val and *HNF1B* -p.Arg303His, but the phenotype of tubulointerstitial kidney disease, hypomagnesemia and pancreas dysfunction was consistent only with *HNF1B*. Typical signs of *PTH1R* mutations such as Murk Jansen type of metaphyseal chondrodysplasia, characterized by abnormal height, hypercalcemia, bone deformities, and renal calcification, were absent (31), and the *PTH1R* p.Ala72Val was carried by the proband’s asymptomatic cousin (Case AIII-3). Using data from our research population cohort, we show that *HNF1B-*p.Arg303His carriers had eGFR and serum magnesium levels that were comparable or lower than those seen in 17q12 microdeletion carriers. Even though the proband’s sibling does not presently have CKD as defined as eGFR < 60 ml/min/1.73m^2^, her eGFR measured twice in her early 20s (less than the 1^st^ percentile for her age and sex) and her serum magnesium level at age 21 were significantly lower than noncarriers of pathogenic *HNF1B* variants (mean [95% CI] 1.8 mg/dL vs. 2.3 [2.1, 2.5], n = 174 individuals measured at the same age in noncarriers). Notably, we observed that 3 of the 4 *HNF1B* -p.Arg303His cases had elevated serum lipase compared to carriers of 17q12 microdeletion and noncarriers of pathogenic *HNF1B* variants. With *HNF1B* as the candidate gene, we further confirmed that the proband and her parent had hypermagnesuria (FEMg) as seen with *HNF1B* extrarenal abnormalities(32, 33).

Our study exemplifies the important role of large, unselected cohorts with robust EHR data to provide corroborating evidence of clinical traits for the gene-disease pair associated with VUS. It is important to note that variable penetrance of monogenic disorders (e.g. *HNF1B)* can make determination of pathogenicity more challenging, and large cohorts can be very useful to provide confidence on pathogenicity (Mirshahi2022-medrxiv). In a disease with high heterogeneity even within the same family, the presence of similar clinical spectrum between the proband, her family members, and the MyCode participant with *HNF1B*-p.Arg303His is highly supportive of this variant being causal for her CKD. Reports of other renal abnormalities in individuals with *HNF1B* p.Arg303Ser and p.Arg303Cys lend support that the arginine at this locus is important in *HNF1B* function. Indeed, this amino acid in the POU_H_ domain is conserved in multiple species including human, mouse, rat, frog, and zebrafish (Supplement Figure 2).

*PKHD1* and *FXYD2* are known transcriptional targets of *HNF1B*, and disturbed transcription of these genes may cause kidney malformation and hypomagnesemia, respectively (34-36). Our luciferase reporter experiments using wild-type *HNF1B* and *HNB1B*-p.Arg303His showed similar transactivation of the *PKHD1* and *FXYD2* promoters. However, absence of an effect by the mutant on transactivation effect does not exclude pathogenicity. For example, *HNF1B*-p.Val61Gly showed comparable transactivation potential to wildtype *HNF1B* in a luciferase reporter assay(37), even though this variant was observed in 3 children with *HNF1B*-related disorders: a child with a single ovary, a single kidney, and a hemi-uterus(7), and a child with prune belly syndrome and congenital genitourinary malformation(37), and a child with multicystic dysplastic kidney(38). Mild to no alterations in *in vitro* studies were also observed in another homeodomain variant, *HNF1B*-p.Arg295His; this variant was observed in a family of multiple kidney and pancreatic anomalies(39, 40). It was previously shown that the C-terminal domain of *HNF1B* and coactivators modify histone acetylase activity(35), hence mutations of *HNF1B* may reduce histone acetylation on target promoters(41). The absent or mild impact of mutants on luciferase activity may be due to transient nature of the expression of *HNF1B* in the assay as well as differences in the chromatin structure(42).

Our study is not without limitations. While clinical features of *HNF1B* were observed in the proband’s grandparent and grand-relative we lacked genetic testing on these 2 individuals(9). In addition, we cannot exclude the presence of unknown pathogenic structural variants in the intronic regions not covered on the gene panel. Regardless, the clinical spectrum observed in the four Arg303His families, the Arg303Ser family, and the Arg303Cys patient provides robust evidence that this locus does not tolerate these amino acid changes.

Despite the high utility of genetic testing in CKD, upwards of 10-30% of tests returned a VUS in part because the painstaking work of gathering and adjudicating evidence to determine the pathogenicity (3, 16-18). Variant classification for CKD-associated variants is disproportionately affected as very few CKD genes are recommended as returnable in incidental findings by the ACMG-AMP guidelines compared to cardiac disease or cancer genes(43). A VUS presents an ethical challenge to report to patients and their families due to insurance liability, genetic counseling availability, and the concern that the disease causality of these variants can be overstated leading to unnecessary stress(44). In families with genetic kidney disease, resolution of VUS is particularly important because detection of genetic kidney disease in family members can not only allow for appropriate precision management but also help avoid a situation where an asymptomatic family member with a genetic pathogenic variant donate a kidney and then later develops ESKD. Similarly, an unresolved VUS could delay family member carriers from serving as potential kidney donors if the variant is later unequivocally determined benign.

In conclusion, we present a VUS observed in *HNF1B* from clinical genetic testing of a kidney transplant candidate and two affected family members. Literature search alone showed that variations at this locus led to kidney disease phenotype consistent with the *HNF1B* disease spectrum; however, there was not enough evidence for pathogenicity. Using a population cohort with genetic and clinical data, we propose a multi-stage process in evaluating VUS. Altogether, the data support the pathogenicity of the *HNF1B*-p.Arg303His variant, providing a genetic diagnosis for the proband and her parent. The proband’s presently asymptomatic sibling can be safely excluded as a possible living donor since she also harbors the same pathogenic variant. The utility of this model can be applied to other genetic diseases as genetic testing become more routine.

## Data Availability

All data produced in the present study are available upon reasonable request to the authors.

## Funding

This work was financially supported by a grant from the Dutch Kidney Foundation to J. de Baaij (Large Kolff grant 17OKG07) and J. Hoenderop (IMAGEN project which is co-funded by the PPP Allowance made available by Health∼Holland, Top Sector Life Sciences & Health, LSHM20009).

## Acknowledgements

The authors would like to acknowledge the participants of the MyCode® Community Health Initiative for use of their genomic and electronic health information, without whom this study would not be possible. The patient enrollment and exome sequencing were funded by the Regeneron Genetics Center. We thank the Geisinger-Regeneron DiscovEHR Collaboration for making the genotype data and phenotype available for this project. We thank Dr. Matthew Oetjens for helpful discussions on the methodology to confirm exome CNV calls using genotyping array. We thank the Maturity-Onset Diabetes of the Young (MODY) Expert Panel for helpful discussions regarding this variant.

## Table of Content

3. Supplemental Table 2. Pathogenic classification for HNF1B Arg303His using ACMG-AMP guidelines.

---

**Supplemental Table 1.** Rare variants among members of Family. **A**. All individuals harbor a single allele (heterozygous state) in each variant noted.

|  | Gene | Variant location | Inheritance pattern | gnomAD MAF (NFE) | Current ClinVar Classification | Disease(s) Associated with Gene | Kidney Condition Observed (Inheritance pattern) [Reference]<sup>s</sup> |
| --- | --- | --- | --- | --- | --- | --- | --- |
| **Case AIII-1 (proband)** | HNF1B | NM_00458.4:c.908G>A (p.Arg303His) | AD | NA | VUS | <u>Renal cysts and diabetes syndrome</u> | Isolated CAKUT (AD)[1]; RCAD (AD)[1] |
|  | TP63 | NM_003722.4:c.1459C>T (p.Arg487Cys) | AD | NA | VUS | Ectrodactyly, ectodermal dysplasia, clefting (EEC) syndrome; acro-dermatoungual-lacrima-tooth (ADULT) syndrome; and limb-mammary syndrome (LMS) | Syndromic CAKUT (AD)[2] |
|  | PTH1R | NM_000316.3:c.215C>T (p.Ala72Val) | AD and AR | 2.6e-5 * | VUS | Chondrodysplasia, Blomstrand Type and Metaphyseal Chondrodysplasia, Jansen Type |  |
|  | FREM1 | NM_144965.5:c.1579A>G (p.Asn527Asp) | AD | 6.6e-6* | VUS | Bifid nose, renal agenesis, and anorectal malformations syndrome, sometimes called BNAR | Isolated CAKUT(AR)[3] |
|  | LRP2 | NM_004525.2:c.6160G>A (p.Asp2054Asn) | AR | 9.7e-4 | VUS | Donnai-Barrow syndrome | Syndromic CAKUT (AR)[4] |
|  | SLC12A2 | NM_001046.2:c.235A>C (p.Ser79Arg) | Unknown | 6.6e-6* | VUS | <u>Deafness, Autosomal Dominant 78 and Delpire-Mcneill Syndrome.</u> |  |
|  | CFTR | NM_000492.4:c.3194T>C (p.Leu1065Pro) | AR |  | Pathogenic | Cystic fibrosis |  |
| **Case AII-1** | HNF1B | NM_00458.4:c.908G>A (p.Arg303His) | AD | NA | VUS | Renal cysts and diabetes syndrome | Isolated CAKUT (AD)[1]; RCAD (AD)[1] |
|  | PKD1 | NM_001009944.3:c.5307T>G (p.His1769Gln) | AD | 1.3e-5* | VUS | Polycystic kidney disease (ADPKD) | Syndromic CAKUT (AD)[5] |
|  | PTH1R | NM_000316.3:c.215C>T (p.Ala72Val) | AD | 2.6e-5 * | VUS | Chondrodysplasia, Blomstrand Type and Metaphyseal Chondrodysplasia, Jansen Type |  |
|  | ALMS1 | NM_015120.4:c.7809C>G (p.Phe2603Leu) | AR | 4.4e-5 | VUS | Alstrom Syndrome | CKD, nephronophthisis (AR)[6] |
|  | ALMS1 | NM_015120.4:c.8071A>G (p.Lys2691Glu) | AR | NA | VUS | Alstrom Syndrome | CKD, nephronophthisis (AR)[6] |
|  | CEP164 | NM_014956.5:c.2744G>A (p.Arg915His) | AR | 2.4e-4 | VUS | Nephronophthisis and Senior-Loken Syndrome | CKD, nephronophthisis (AR)[7] |
|  | PRODH | NM_016335.6:c.1322T>C (p.Leu441Pro) | AR | 1.9e-3 | VUS | Schizophrenia through linkage, association, and the 22q11 deletion syndrome (Velo-Cardio-Facial syndrome) |  |
|  | SLC12A2 | NM_001046.2:c.235A>C (p.Ser79Arg) | Unknown | 6.6e-6* | VUS | <u>Deafness, Autosomal Dominant 78 and Delpire-Mcneill Syndrome.</u> |  |
|  | CFTR | NM_000492.4:c.3194T>C (p.Leu1065Pro) | AR |  | Pathogenic | Cystic fibrosis |  |
| **Case AIII-2** | HNF1B | NM_00458.4:c.908G>A (p.Arg303His) | AD | NA | VUS | Renal cysts and diabetes syndrome | Isolated CAKUT (AD)[1]; RCAD (AD)[1] |
|  | TP63 | NM_003722.4:c.1459C>T<br>(p.Arg487Cys) | AD | NA | VUS | Ectrodactyly, ectodermal dysplasia, clefting (EEC) syndrome; acro-dermato-ungual-lacrima-tooth (ADULT) syndrome; and limb-mammary syndrome (LMS) | Isolated CAKUT (AD)[2] |
|  | PKD1 | NM_001009944.3:c.5307T>G<br>(p.His1769Gln) | AD | 1.3e-5* | VUS | Polycystic kidney disease (ADPKD) | Syndromic CAKUT (AD)[5] |
|  | PTH1R | NM_000316.3:c.215C>T<br>(p.Ala72Val) | AD and AR | 2.6e-5 * | VUS | Chondrodysplasia, Blomstrand Type and Metaphyseal Chondrodysplasia, Jansen Type |  |
| **Case AII-2** | NPHS2 | NM_014625.4:c.686G>A<br>(p.Arg229Gln) | AR | NA | VUS | Nephrotic syndrome, type 2 | Nephrotic syndrome (AR)[8] |
| **Case AIII-3** | NPHS2 | NM_014625.4:c.686G>A<br>(p.Arg229Gln) | AR | NA | VUS | Nephrotic syndrome, type 2 | Nephrotic syndrome (AR)[8] |
|  | PTH1R | NM_000316.3:c.215C>T<br>(p.Ala72Val) | AD and AR | 2.6e-5 * | VUS | Chondrodysplasia, Blomstrand Type and Metaphyseal Chondrodysplasia, Jansen Type |  |
|  | CFI | NM_000204.5:c.1311dup<br>(p.Asp438Argfs*8) | AD and AR | 6.57e-6 * |  | Complement factor I deficiency; Hemolytic uremic syndrome, atypical; Macular degeneration, age-related | Chronic glomerulonephritis (AR)[9] |

\*not in non-Finnish European (NFE)
§As reviewed by Connaughton *et al*<sup>10</sup>
Abbreviations: AD autosomal dominant, AR autosomal recessive, MAF minor allele frequency, VUS variant of unknown significance

**Supplemental Figure 1.**
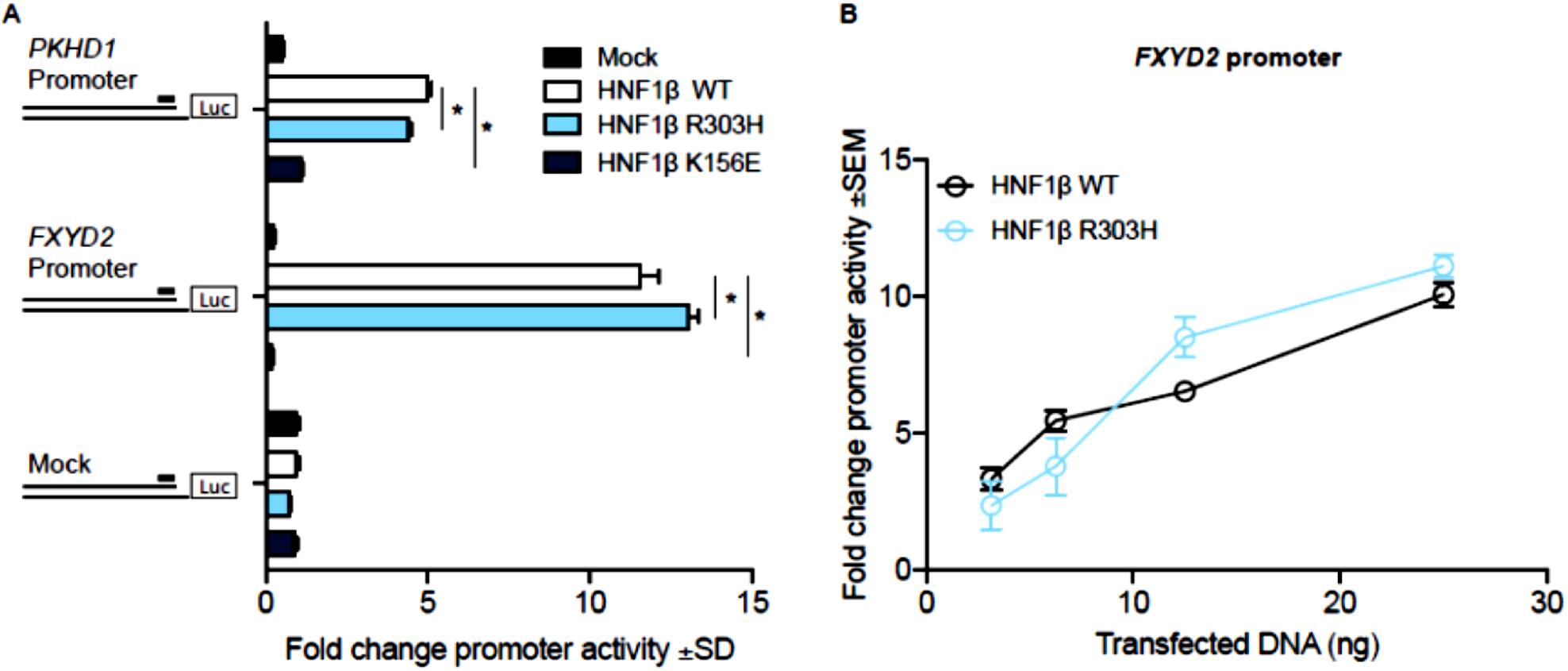
*In vitro* experiments showed mildly altered, differential transactivation potential of HNF1B-p.Arg303His. **A**. HEK 293 cells were transiently transfected with 350 ng of a luciferase construct carrying the human *FXYD2, PKHD1* promoter, or an empty vector mock construct. Promoter activity was tested in the presence of 25 ng of a construct carrying wildtype *HNF1BHNF1B, HNF1B*-p.Arg303His, positive control *HNF1B*-p.Lys156Glu, or a mock construct. Firefly luciferase signal were normalized to luminescence from Renilla luciferase-CMV promoter for each condition in 3 determinations. Fold change promoter activity compared to mock Luciferase/mock Renilla luminescence + SD are shown. **B**. Saturation curve of luciferase transcriptional activity of wildtype *HNF1B* and *HNF1B-*p.Arg303His on *FXYD2* promoter as assessed in increasing amounts of transfected DNA constructs. HEK 293 cells were co-transfected with a luciferase construct carrying the human *FXYD2* promoter and wildtype *HNF1B* or *HNF1B-*p.R303H at amounts indicated. Luciferase activity is expressed as fold change promoter activity compared to mock/mock luminescence. Values are mean + SEM in triplicate determinations. **C**. Same as B except for *PKHD1* promoter. *p<0.05, two-way Analysis of Variance (ANOVA).

**Supplemental Figure 2.**
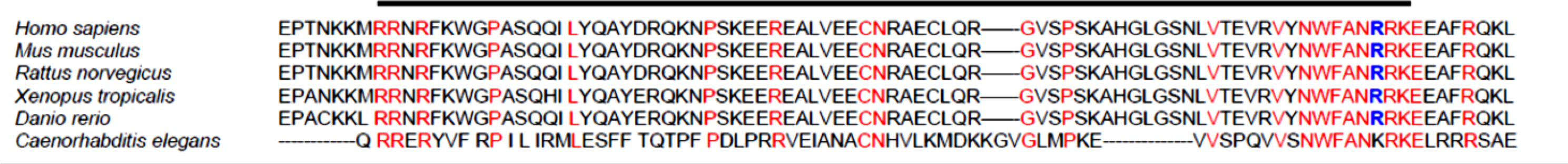
The Arg303 is conserved in the POU homeodomain of *HNF1B*. Protein sequence alignment of the POU_H_ domain (amino acids 232 to 305 in human, highlighted by the black bar) in human, mouse, rat, xenopus, zebrafish, and roundworm adapted from MARRVEL multiple protein alignment tool ^11^. Conserved amino acids among all species represented are in red. Note that all amino acids of the POU_H_ domain are conserved in vertebrates, including arg303, which is highlighted in bold blue.

## Notes

### Competing Interest Statement

The authors have declared no competing interest.

### Author Declarations

Ethics committee/IRB of Geisinger Medical Center gave ethical approval for this work.

## Reference

1. Westland R, Renkema KY, Knoers N. Clinical Integration of Genome Diagnostics for Congenital Anomalies of the Kidney and Urinary Tract. Clin J Am Soc Nephrol. 2020;16(1):128–37.

2. Groopman EE, Marasa M, Cameron-Christie S, Petrovski S, Aggarwal VS, Milo-Rasouly H, et al. Diagnostic Utility of Exome Sequencing for Kidney Disease. N Engl J Med. 2019;380(2):142–51.

3. Connaughton DM, Kennedy C, Shril S, Mann N, Murray SL, Williams PA, et al. Monogenic causes of chronic kidney disease in adults. Kidney Int. 2019;95(4):914–28.

4. Mann N, Braun DA, Amann K, Tan W, Shril S, Connaughton DM, et al. Whole-Exome Sequencing Enables a Precision Medicine Approach for Kidney Transplant Recipients. J Am Soc Nephrol. 2019;30(2):201–15.

5. Knoers N, Antignac C, Bergmann C, Dahan K, Giglio S, Heidet L, et al. Genetic testing in the diagnosis of chronic kidney disease: recommendations for clinical practice. Nephrol Dial Transplant. 2022;37(2):239–54.

6. Jayasinghe K, Stark Z, Kerr PG, Gaff C, Martyn M, Whitlam J, et al. Clinical impact of genomic testing in patients with suspected monogenic kidney disease. Genet Med. 2021;23(1):183–91.

7. Edghill EL, Bingham C, Ellard S, Hattersley AT. Mutations in hepatocyte nuclear factor-1beta and their related phenotypes. J Med Genet. 432006. p. 84–90.

8. Ulinski T, Lescure S, Beaufils S, Guigonis V, Decramer S, Morin D, et al. Renal phenotypes related to hepatocyte nuclear factor-1beta (TCF2) mutations in a pediatric cohort. J Am Soc Nephrol. 2006;17(2):497–503.

9. Clissold RL, Hamilton AJ, Hattersley AT, Ellard S, Bingham C. HNF1B-associated renal and extra-renal disease-an expanding clinical spectrum. Nat Rev Nephrol. 2015;11(2):102–12.

10. Adalat S, Woolf AS, Johnstone KA, Wirsing A, Harries LW, Long DA, et al. HNF1B mutations associate with hypomagnesemia and renal magnesium wasting. J Am Soc Nephrol. 2009;20(5):1123–31.

11. Mallett AJ, McCarthy HJ, Ho G, Holman K, Farnsworth E, Patel C, et al. Massively parallel sequencing and targeted exomes in familial kidney disease can diagnose underlying genetic disorders. Kidney Int. 2017;92(6):1493–506.

12. Rao J, Liu X, Mao J, Tang X, Shen Q, Li G, et al. Genetic spectrum of renal disease for 1001 Chinese children based on a multicenter registration system. Clin Genet. 2019.

13. Eckardt KU, Alper SL, Antignac C, Bleyer AJ, Chauveau D, Dahan K, et al. Autosomal dominant tubulointerstitial kidney disease: diagnosis, classification, and management--A KDIGO consensus report. Kidney Int. 2015;88(4):676–83.

14. Cocchi E, Nestor JG, Gharavi AG. Clinical Genetic Screening in Adult Patients with Kidney Disease. Clin J Am Soc Nephrol. 2020;15(10):1497–510.

15. Phillips KA, Deverka PA, Hooker GW, Douglas MP. Genetic Test Availability And Spending: Where Are We Now? Where Are We Going? Health Aff (Millwood). 2018;37(5):710–6.

16. Elhassan EAE, Murray SL, Connaughton DM, Kennedy C, Cormican S, Cowhig C, et al. The utility of a genetic kidney disease clinic employing a broad range of genomic testing platforms: experience of the Irish Kidney Gene Project. J Nephrol. 2022.

17. Thomas CP, Freese ME, Ounda A, Jetton JG, Holida M, Noureddine L, et al. Initial experience from a renal genetics clinic demonstrates a distinct role in patient management. Genet Med. 2020;22(6):1025–35.

18. Lieberman KV, Chang AR, Block GA, Robinson K, Bristow SL, Devarajan P, et al. The KIDNEYCODE program: Diagnostic yield and clinical features of individuals with chronic kidney disease. Kidney360. 2022:10.34067/KID.0004162021.

19. Hay E, Cullup T, Barnicoat A. A practical approach to the genomics of kidney disorders. Pediatr Nephrol. 2022;37(1):21–35.

20. Carey DJ, Fetterolf SN, Davis FD, Faucett WA, Kirchner HL, Mirshahi U, et al. The Geisinger MyCode community health initiative: an electronic health record-linked biobank for precision medicine research. Genet Med. 2016;18(9):906–13.

21. Mirshahi UL, Luo JZ, Manickam K, Wardeh AH, Mirshahi T, Murray MF, et al. Trajectory of exonic variant discovery in a large clinical population: implications for variant curation. Genet Med. 2019;21(6):1417–24.

22. Richards S, Aziz N, Bale S, Bick D, Das S, Gastier-Foster J, et al. Standards and guidelines for the interpretation of sequence variants: a joint consensus recommendation of the American College of Medical Genetics and Genomics and the Association for Molecular Pathology. Genet Med. 2015;17(5):405–24.

23. Jarvik GP, Browning BL. Consideration of Cosegregation in the Pathogenicity Classification of Genomic Variants. Am J Hum Genet. 2016;98(6):1077–81.

24. Packer JS, Maxwell EK, O’Dushlaine C, Lopez AE, Dewey FE, Chernomorsky R, et al. CLAMMS: a scalable algorithm for calling common and rare copy number variants from exome sequencing data. Bioinformatics. 2016;32(1):133–5.

25. Fortier N, Rudy G, Scherer A. Detection of CNVs in NGS Data Using VS-CNV. Methods Mol Biol. 2018;1833:115–27.

26. Landrum MJ, Lee JM, Benson M, Brown GR, Chao C, Chitipiralla S, et al. ClinVar: improving access to variant interpretations and supporting evidence. Nucleic Acids Res. 2018;46(D1):D1062–d7.

27. Kompatscher A, de Baaij JHF, Aboudehen K, Hoefnagels A, Igarashi P, Bindels RJM, et al. Loss of transcriptional activation of the potassium channel Kir5.1 by HNF1β drives autosomal dominant tubulointerstitial kidney disease. Kidney Int. 2017;92(5):1145–56.

28. Schwartz GJ, Muñoz A, Schneider MF, Mak RH, Kaskel F, Warady BA, et al. New equations to estimate GFR in children with CKD. J Am Soc Nephrol. 2009;20(3):629–37.

29. Levey AS, Stevens LA, Schmid CH, Zhang YL, Castro AF, 3rd, Feldman HI, et al. A new equation to estimate glomerular filtration rate. Annals of Internal Medicine. 2009;150(9):604–12.

30. Snoek R, Nguyen TQ, van der Zwaag B, van Zuilen AD, Kruis HME, van Gils-Verrij LA, et al. Importance of Genetic Diagnostics in Adult-Onset Focal Segmental Glomerulosclerosis. Nephron. 142: © 2019 The Author(s) Published by S. Karger AG, Basel.; 2019. p. 351–8.

31. Saito H, Noda H, Gatault P, Bockenhauer D, Loke KY, Hiort O, et al. Progression of Mineral Ion Abnormalities in Patients With Jansen Metaphyseal Chondrodysplasia. J Clin Endocrinol Metab. 2018;103(7):2660–9.

32. Adalat S, Hayes WN, Bryant WA, Booth J, Woolf AS, Kleta R, et al. HNF1B Mutations Are Associated With a Gitelman-like Tubulopathy That Develops During Childhood. Kidney Int Rep. 2019;4(9):1304–11.

33. van der Made CI, Hoorn EJ, de la Faille R, Karaaslan H, Knoers NV, Hoenderop JG, et al. Hypomagnesemia as First Clinical Manifestation of ADTKD-HNF1B: A Case Series and Literature Review. Am J Nephrol. 2015;42(1):85–90.

34. Ferrè S, de Baaij JH, Ferreira P, Germann R, de Klerk JB, Lavrijsen M, et al. Mutations in PCBD1 cause hypomagnesemia and renal magnesium wasting. J Am Soc Nephrol. 2014;25(3):574–86.

35. Hiesberger T, Shao X, Gourley E, Reimann A, Pontoglio M, Igarashi P. Role of the hepatocyte nuclear factor-1beta (HNF-1beta) C-terminal domain in Pkhd1 (ARPKD) gene transcription and renal cystogenesis. J Biol Chem. 2005;280(11):10578–86.

36. Ferrè S, Veenstra GJ, Bouwmeester R, Hoenderop JG, Bindels RJ. HNF-1B specifically regulates the transcription of the γa-subunit of the Na+/K+-ATPase. Biochem Biophys Res Commun. 2011;404(1):284–90.

37. Granberg CF, Harrison SM, Dajusta D, Zhang S, Hajarnis S, Igarashi P, et al. Genetic basis of prune belly syndrome: screening for HNF1β gene. J Urol. 2012;187(1):272–8.

38. Hoskins BE, Cramer CH, 2nd, Tasic V, Kehinde EO, Ashraf S, Bogdanovic R, et al. Missense mutations in EYA1 and TCF2 are a rare cause of urinary tract malformations. Nephrol Dial Transplant. 23. England2008. p. 777–9.

39. Bohn S, Thomas H, Turan G, Ellard S, Bingham C, Hattersley AT, et al. Distinct molecular and morphogenetic properties of mutations in the human HNF1beta gene that lead to defective kidney development. J Am Soc Nephrol. 2003;14(8):2033–41.

40. Barbacci E, Chalkiadaki A, Masdeu C, Haumaitre C, Lokmane L, Loirat C, et al. HNF1beta/TCF2 mutations impair transactivation potential through altered co-regulator recruitment. Hum Mol Genet. 2004;13(24):3139–49.

41. Gong Y, Ma Z, Patel V, Fischer E, Hiesberger T, Pontoglio M, et al. HNF-1beta regulates transcription of the PKD modifier gene Kif12. J Am Soc Nephrol. 2009;20(1):41–7.

42. Inoue F, Kircher M, Martin B, Cooper GM, Witten DM, McManus MT, et al. A systematic comparison reveals substantial differences in chromosomal versus episomal encoding of enhancer activity. Genome Res. 2017;27(1):38–52.

43. Green RC, Berg JS, Grody WW, Kalia SS, Korf BR, Martin CL, et al. ACMG recommendations for reporting of incidental findings in clinical exome and genome sequencing. Genet Med. 2013;15(7):565–74.

44. Savatt JM, Myers SM. Genetic Testing in Neurodevelopmental Disorders. Front Pediatr. 2021;9:526779.

## References

1. Horikawa Y, Iwasaki N, Hara M, et al. Mutation in hepatocyte nuclear factor-1 beta gene (TCF2) associated with MODY. Nat Genet. 1997;17(4):384–385. Accessed Mar 18, 2022. doi: 10.1038/ng1297-384.

2. Celli J, Duijf P, Hamel BC, et al. Heterozygous germline mutations in the p53 homolog p63 are the cause of EEC syndrome. Cell. 1999;99(2):143–153. doi: S0092-8674(00)81646-3 [pii].

3. Kohl S, Hwang D, Dworschak GC, et al. Mild recessive mutations in six fraser syndrome-related genes cause isolated congenital anomalies of the kidney and urinary tract. J Am Soc Nephrol. 2014;25(9):1917–1922. Accessed Mar 18, 2022. doi: 10.1681/ASN.2013101103.

4. Kantarci S, Al-Gazali L, Hill RS, et al. Mutations in LRP2, which encodes the multiligand receptor megalin, cause donnai-barrow and facio-oculo-acoustico-renal syndromes. Nat Genet. 2007;39(8):957–959. Accessed Mar 18, 2022. doi: 10.1038/ng2063.

5. Rossetti S, Consugar MB, Chapman AB, et al. Comprehensive molecular diagnostics in autosomal dominant polycystic kidney disease. J Am Soc Nephrol. 2007;18(7):2143–2160. Accessed Mar 18, 2022. doi: 10.1681/ASN.2006121387.

6. Collin GB, Marshall JD, Ikeda A, et al. Mutations in ALMS1 cause obesity, type 2 diabetes and neurosensory degeneration in alström syndrome. Nat Genet. 2002;31(1):74–78. Accessed Mar 18, 2022. doi: 10.1038/ng867.

7. Chaki M, Airik R, Ghosh AK, et al. Exome capture reveals ZNF423 and CEP164 mutations, linking renal ciliopathies to DNA damage response signaling. Cell. 2012;150(3):533–548. Accessed Mar 18, 2022. doi: 10.1016/j.cell.2012.06.028.

8. Boute N, Gribouval O, Roselli S, et al. NPHS2, encoding the glomerular protein podocin, is mutated in autosomal recessive steroid-resistant nephrotic syndrome. Nat Genet. 2000;24(4):349–354. Accessed Mar 18, 2022. doi: 10.1038/74166.

9. Fremeaux-Bacchi V, Dragon-Durey M-, Blouin J, et al. Complement factor I: A susceptibility gene for atypical haemolytic uraemic syndrome. J Med Genet. 2004;41(6):e84. Accessed Mar 18, 2022. doi: 10.1136/jmg.2004.019083.

10. Connaughton DM, Kennedy C, Shril S, et al. Monogenic causes of chronic kidney disease in adults. Kidney Int. 2019;95(4):914–928. Accessed Mar 18, 2022. doi: 10.1016/j.kint.2018.10.031.

11. Wang J, Al-Ouran R, Hu Y, et al. MARRVEL: Integration of human and model organism genetic resources to facilitate functional annotation of the human genome. Am J Hum Genet. 2017;100(6):843–853. Accessed Mar 18, 2022. doi: 10.1016/j.ajhg.2017.04.010.

